# Evaluating Artificial Intelligence Assisted Nursing Education: Student Perceptions, Ethical Concerns, and Pedagogical Implications

**DOI:** 10.1101/2025.06.09.25329272

**Authors:** Ramteja Sajja, Yusuf Sermet, Ibrahim Demir

## Abstract

**Background:** Artificial intelligence (AI) tools are increasingly being integrated into nursing education to enhance learning and provide flexible academic assistance. However, little is known about how undergraduate nursing students perceive these tools or how they affect learning experiences.

**Aim:** To evaluate nursing students’ perceptions of an AI-powered academic assistant and assess its perceived usefulness, trustworthiness, and ethical implications within a real course setting.

**Methods:** This quantitative study was conducted in a junior-level undergraduate nursing course at a large public university. Students (N = 38) completed pre- and post-surveys measuring their attitudes, confidence, ethical concerns, and engagement with the Educational AI Hub. System usage data were also analyzed to assess tool interaction.

**Results:** Students reported high levels of convenience and comfort using the AI tool, particularly for studying and concept review. However, concerns emerged around academic integrity and uncertainty about appropriate use. Most students supported moderate restrictions and expressed strong interest in future AI integration.

**Conclusions:** AI tools can support associate degree nursing students by enhancing independent study and access to learning support. Clear guidelines and ethical frameworks are essential for responsible implementation.

## 1. Introduction

Artificial intelligence (AI) is rapidly reshaping healthcare and nursing education, offering transformative possibilities for how students learn, engage, and prepare for clinical practice in a digitally evolving healthcare landscape (Srinivasan et al., 2024; Tam et al., 2023). AI-enhanced technologies such as intelligent chatbots, adaptive quizzes, and virtual simulations are increasingly being integrated into academic settings to improve learning outcomes and streamline instruction (Zhang et al., 2023; Castonguay et al., 2023). Additionally, tools for automated content generation and self-assessment are fostering more self-directed and personalized learning experiences for students (Jallad et al., 2024; Tseng et al., 2025; Labrague & Sabei, 2024). These innovations signal a significant shift in educational strategies and pedagogical approaches across health professions education (Sermet & Demir, 2021).

This shift aligns with broader educational movements toward competency-based, student-centered, and digitally mediated learning, where AI enables more flexible, personalized, and skills-focused instruction. AI-driven platforms support competency-based education by adapting content and assessments to individual learner needs, helping students progress at their own pace while achieving clearly defined outcomes (Ghnemat et al., 2022; Krive et al., 2023; Mady & Niese, 2022). They also enhance personalization by identifying learning gaps early and adjusting pathways accordingly, which fosters self-regulation and supports diverse learning profiles (Msambwa et al., 2025; Bhutoria, 2022). In addition, AI technologies have been shown to improve student engagement and reflective thinking through real-world simulations, collaborative tasks, and inquiry-based learning approaches (Saritepeci & Durak, 2024; Kit et al., 2022; Ghnemat et al., 2022). These tools not only transform the student experience but also require educators to adopt new digital and AI-related competencies to ethically and effectively guide learners in this evolving landscape (Ng et al., 2023; Xu & Fan, 2021; Bhutoria, 2022).

Building on this foundation, AI tools offer a wide spectrum of educational benefits, ranging from curriculum support (Pursnani et al., 2023) to professional training (Sajja et al., 2025a; Pursnani et al., 2025). Personalized learning through platforms that include chatbots, flashcards, and adaptive assessments allows students to receive immediate feedback, revisit difficult content, and study at their own pace (Charow et al., 2021; Baglivo et al., 2023; Sapci & Sapci, 2020). These features support diverse learning styles and contribute to increased content mastery (Zhang et al., 2023; Balay-Odao et al., 2024). Advanced simulations and AI-driven virtual reality environments create authentic, risk-free environments in which students can practice clinical judgment, interprofessional communication, and complex decision-making (Jallad et al., 2024; Montejo et al., 2024; Labrague & Sabei, 2024). These tools are further complemented by generative AI technologies like ChatGPT, which assist in the preparation of patient education materials, clinical case analyses, and academic writing, resulting in improved clarity, actionability, and quality (Saatçi et al., 2024; Silvestri-Elmore & Burton, 2024; Tseng et al., 2025).

Emerging frameworks such as the AI-enabled Intelligent Assistant (AIIA) and virtual teaching assistants based on GPT-3 further exemplify the potential of AI to provide scalable, course-specific, and interactive support for students, promoting autonomous learning while alleviating instructor workload (Sajja et al., 2023a; Sajja et al., 2024). These systems offer capabilities such as generating quizzes, answering logistical and content-related questions, and crafting individualized learning pathways. In tandem, recent advancements in learning analytics tools powered by GPT-4 have enabled educators to quantify student engagement, map cognitive development through Bloom’s taxonomy, and make data-informed instructional decisions, though challenges around data security and interpretability remain (Sajja et al., 2023b). Moreover, domain-specific fine-tuning of semantic search models has enhanced educational chatbots’ ability to understand academic language and retrieve relevant information from learning management systems and course materials (Sajja et al., 2025c). Similar approaches have been successfully applied in other disciplines such as hydrology, where benchmarking tools like HydroLLM-Benchmark have demonstrated the value of assessing LLM performance within specialized fields (Kizilkaya et al., 2025). These developments, though not specific to nursing, provide foundational architectures that can be adapted to healthcare education environments (Abrar et al., 2025).

Beyond enhancing knowledge and technical skills, AI tools have demonstrated potential to foster effective and behavioral competencies essential to professional nursing practice. Personalized, gamified, and interactive AI and web systems increase motivation and engagement, while cultivating autonomy and digital literacy, skills necessary for lifelong learning and adaptability in technology-driven clinical environments (Chi et al., 2025; Srinivasan et al., 2024; Kowitlawakul et al., 2022). These platforms also promote learner persistence and engagement through adaptive content delivery and real-time feedback mechanisms (Labrague et al., 2023; Younas et al., 2025; Sukmawati et al., 2025). Furthermore, student attitudes toward AI’s utility in healthcare practice are positively correlated with their willingness to engage meaningfully with these tools in academic and clinical contexts (Labrague et al., 2023; Kwak et al., 2022).

Yet, integrating AI into nursing education also introduces notable challenges. Ethical concerns about academic integrity, over-reliance, plagiarism, and data privacy persist (Topaz et al., 2024; Zohny et al., 2023; De Gagne et al., 2023). These issues raise important questions about responsible use and the boundaries of academic support tools (Lane et al., 2024; Ali & Aysan, 2024). Students and educators alike must contend with fairness, transparency, and the risk of misinformation or bias in AI-generated content (Castonguay et al., 2023; Simms, 2024). These risks may undermine trust in educational technologies if not properly addressed (Hawk et al., 2024; Han et al., 2025). Additionally, AI tools may lack the human nuance required to support ethical reflection, empathy, and person-centered care, core values in nursing education. Concerns also remain about the impact of AI on students’ development of critical thinking, clinical reasoning, and independent decision-making, particularly when AI use is not thoughtfully scaffolded (Shin et al., 2024; Benfatah et al., 2024; Alshehri et al., 2022).

This duality in potential, promise and risk, highlight a pressing need for evidence-informed guidance on the integration of AI into nursing education. As nurse educators aim to cultivate not just competency but also empathy, advocacy, and professional engagement, understanding how AI tools influence both cognitive and affective outcomes become imperative. AI cannot be a mere content delivery mechanism; rather, it must be embedded within pedagogies that promote reflective thinking, ethical awareness, and sustained professional identity formation. Given this evolving educational landscape, it is essential to understand how nursing students perceive, interact with, and are impacted by AI-powered tools in authentic academic environments. Recent work in engineering education has shown that while students find AI assistants helpful for learning and task completion, their engagement is significantly influenced by usability, institutional policy clarity, and ethical guidance (Sajja et al., 2025d).

Despite the increasing adoption of AI in health professions education, limited empirical evidence exists on how these tools affect student experience in real-world nursing curricula. This study investigates the implementation of an AI-enhanced learning assistant, Educational AI Hub (Sajja et al., 2025b), within a nursing course, focusing on students’ experiences, usage patterns, and perceived learning outcomes. By examining the intersections of trust, usability, and academic engagement, as well as the ethical dimensions of AI use in nursing education, this research offers insights into the responsible and effective integration of AI to support professional development, critical thinking, and learner empowerment in nursing curricula.

## 2. Methodology

This study employed a quantitative approach to evaluate the use and impact of the Educational AI Hub (Sajja et al., 2025b); an AI-powered learning assistant designed to support nursing students. The methodology focused on capturing a comprehensive understanding of how students interacted with the tool, how they perceived its usefulness, and how it influenced their learning experiences. Key components of the methodology included a detailed overview of the AI tool’s features, participant demographics, and pre- and post-intervention survey instruments. Additionally, system-generated usage data provided insights into how students engaged with functionalities such as the chatbot, flashcards, quizzes, and note-generation tools. This structured approach enabled the measurement of behavioral patterns and shifts in student perceptions within the context of a real nursing course.

### 2.1. Educational AI Hub Features

The Educational AI Hub is an integrated, AI-powered learning assistant developed to support nursing students by offering a suite of intelligent, personalized learning tools. Embedded directly into the course’s learning management system, the tool is designed to enhance student understanding, engagement, and academic performance through six key functionalities.

**Notes Generation:** This feature allows students to enter a topic of interest, and the system responds by producing concise, structured summaries synthesized from reliable academic content. These AI-generated notes help students quickly comprehend complex topics, facilitating efficient review and supporting self-paced learning.

**AI-Assisted Chatbot:** The chatbot offers immediate responses to course-related questions, functioning as a virtual tutor available at any time. It maintains short-term conversational memory to provide contextually relevant answers and adapts its tone based on the user’s interaction style. This feature enhances student engagement and extends academic support beyond traditional classroom boundaries.

**Flashcards Generation:** By submitting a topic, students can generate digital flashcards based on Bloom’s Digital Taxonomy. These cards span a range of cognitive domains, from basic recall to higher-level thinking such as analysis and application, enabling students to reinforce knowledge and engage with material at varying depths.

**Quiz Generation and Grading:** Students can create customized quizzes on course topics to assess their understanding. The quiz items are designed to target different levels of cognitive complexity, aligning with Bloom’s taxonomy. Upon completion, students receive immediate feedback and performance summaries. The system uses a flexible grading algorithm that evaluates the semantic accuracy of responses, allowing for variation in wording while still accurately assessing comprehension. This promotes deeper learning and encourages students to articulate their knowledge in their own words. As illustrated in Figure 1, the Educational AI Hub interface provides an intuitive layout for accessing the quiz feature, enabling efficient quiz creation and interactive feedback delivery.

**Figure 1:**
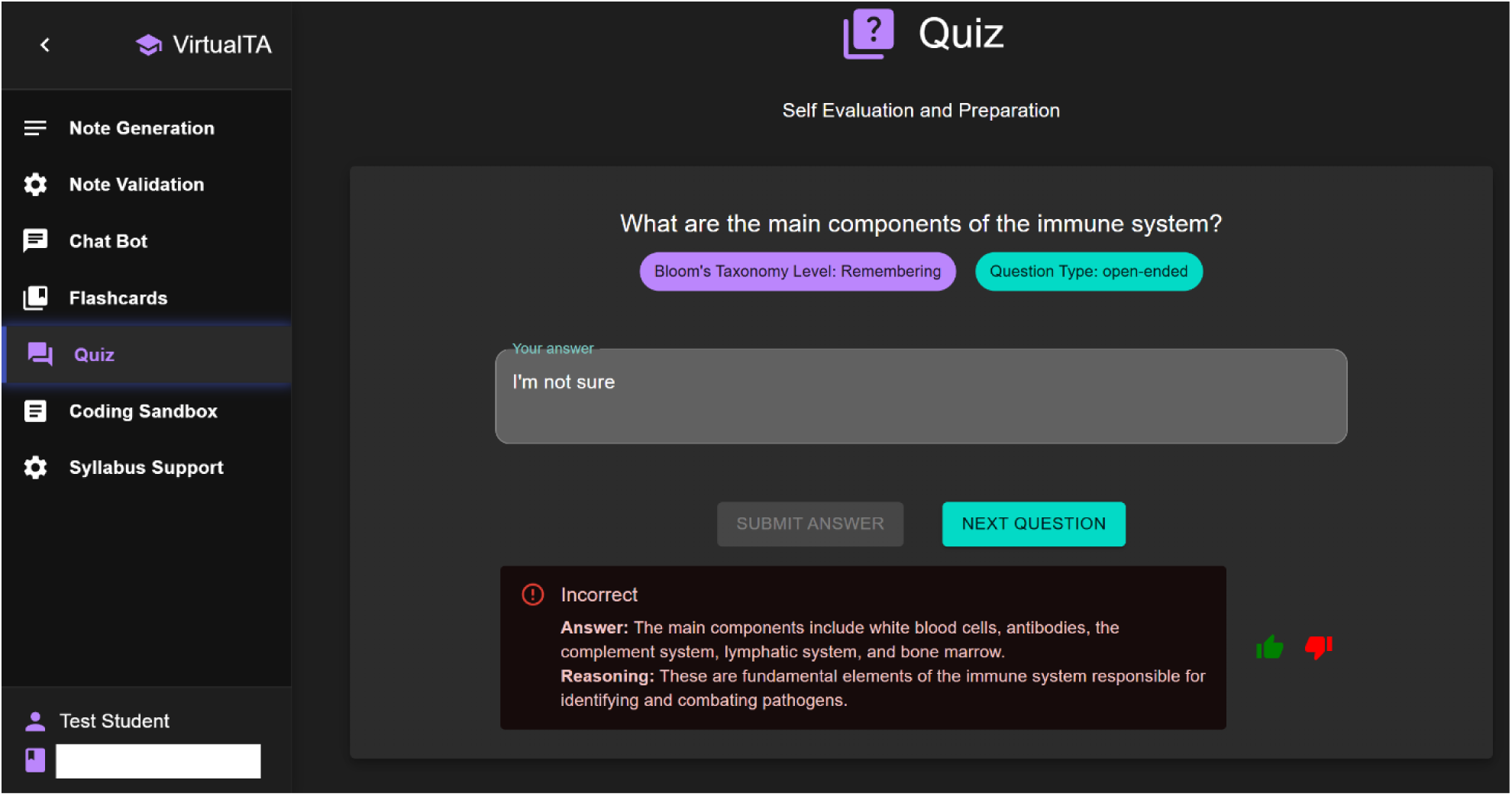
Educational AI Hub Interface showing the quiz feature

**Coding Sandbox:** Although designed primarily for technical learning contexts, this feature supports problem-solving by allowing users to input programming-related questions. The system provides tiered assistance, starting with pseudocode and progressing to full explanations and runnable code, helping students build skills in computational tools where applicable to course content.

**Syllabus Support:** Students can also use the AI assistant to ask logistical and administrative questions related to the course, such as assignment due dates or exam schedules. The system accesses the course syllabus to deliver accurate, timely responses, improving clarity and helping students stay organized.

Collectively, these features offer a flexible, student-centered approach to learning, enabling nursing students to access academic support, reinforce key concepts, and navigate course materials more effectively. While the current study focuses on the Educational AI Hub as a single implementation, the tool’s modular design supports integration with a variety of large language models and AI services. This generalizability positions the Hub not only as a course-specific solution but also as a scalable framework applicable across different educational contexts. Furthermore, its comprehensive scope, incorporating both content-focused and administrative support features, makes it one of the most robust AI-powered educational tools available for empirical evaluation. As such, the findings from this study can inform broader discussions on the efficacy and future design of AI-assisted learning environments in health education and beyond.

### 2.2. Study Design

This study was conducted in a junior-level undergraduate course within the College of Nursing at a large R1 public university in the Midwest. The course was selected due to its emphasis on content mastery, critical thinking, and student engagement, core elements that align well with the intended support provided by AI-powered learning tools. The course setting offered an authentic academic environment in which to evaluate how students interact with and perceive AI-enhanced educational technology in nursing education.

A total of 57 students were enrolled in the course. Of these, 38 students participated in the study, yielding a participation rate of approximately 67%. Participation in the pre- and post-surveys varied, with 34 students completing the pre-survey and 19 students completing the post-survey. Additionally, 19 students actively used the Educational AI Hub at least once during the course.

Among the 38 study participants, 6 identified as male and 32 as female. In terms of academic program, 32 students were pursuing a Bachelor of Science in Nursing, while the remaining students were enrolled in related undergraduate degree paths, including Bachelor of Science (n = 3) and Bachelor of Arts (n = 3). Students also reported a variety of declared majors. The majority (n = 32) were Nursing-RN majors, with others majoring in psychology (n = 2), biology (n = 1), biochemistry and molecular biology (n = 1), public health (n = 1), and political science (n = 1).

Students were introduced to the Educational AI Hub through the course’s learning management system and were given access to its full range of features. The AI Hub was presented as an optional learning resource designed to support self-directed study, enhance content comprehension, and provide academic assistance throughout the term.

To evaluate the impact of the tool, the study employed a quantitative design consisting of two main data sources: (1) pre- and post-intervention surveys measuring students’ attitudes, confidence, and perceived value of the tool; and (2) system-generated usage data, which provided insights into how students engaged with various AI Hub features, including the chatbot, flashcards, quizzes, and note-generation tools. This design enabled an assessment of both user behavior and changes in student perceptions over time, offering a structured view of the Educational AI Hub’s potential within a nursing education context.

### 2.3. Data Collection and Analysis

**Pre-Usage Survey:** At the beginning of the semester, a pre-usage survey was distributed to gather baseline data on students’ familiarity with generative AI and their initial attitudes toward its use. The survey assessed prior experience with AI tools in both academic and everyday contexts, as well as students’ confidence in using AI responsibly. It also explored students’ expectations regarding how AI might influence their academic performance, study habits, and productivity. Additional questions addressed trust in AI-generated content and ethical concerns, including views on fairness and the potential for academic dishonesty when using such tools. Responses were collected using a combination of Likert-scale ratings, multiple-choice items, and a limited number of open-ended prompts. These initial responses provided a reference point for later comparison with post-usage survey data to determine how perceptions evolved after direct interaction with the Educational AI Hub.

**Post-Usage Survey:** At the end of the semester, a follow-up survey was carried out to evaluate students’ experiences using the Educational AI Hub. This instrument measured several key areas: (1) the perceived usefulness and ease of using the tool, including comparisons to traditional support resources like teaching assistants or instructors; (2) the impact of the tool on various academic tasks, such as studying, understanding course material, completing assignments, and overall academic progress; (3) students’ comfort and trust in using AI for learning support; (4) ongoing concerns about ethical use, academic integrity, and fairness; and (5) awareness of institutional guidelines regarding AI use in coursework. The survey included structured Likert-scale questions along with optional open-response fields to allow students to share more detailed feedback based on their actual engagement with the platform.

**System Usage Logs:** Throughout the term, automated logs were collected to capture detailed data on student interactions with the Educational AI Hub. These logs offered objective insights into how frequently and in what ways students engaged with the system’s different features, including the chatbot, note generator, flashcards, quiz creation, and coding support tools. Metrics collected included how often each feature was used, types of queries submitted, timing and duration of sessions, and whether students revisited the tool multiple times. Additionally, the system tracked specific actions, such as downloading materials or following up on generated content, providing a clearer picture of the depth of engagement. These usage data were analyzed quantitatively to identify behavioral trends and were compared against survey responses to explore connections between usage patterns and students’ reported perceptions and learning outcomes.

**Open-Ended Feedback:** At the end of the post-survey, students were invited to provide additional reflections through an optional open-ended question: “If you would like, please share any final comments you have related to your experiences with generative AI in this course.” While responses to this prompt were not analyzed in the current study, the question was included to give students an opportunity to express further insights. Instead, for qualitative context, the study focused on the categorized content of queries submitted directly to the AI system during tool use.

**Recruitment and Ethical Considerations:** Participants were recruited via course announcements posted on the learning management system at the beginning of the semester. To encourage participation, students were offered a small amount of extra credit for completing both the pre- and post-surveys. Participation was voluntary, and students who chose not to engage with the AI tools or surveys were allowed to complete a reflective writing assignment for equal credit. All study procedures, including recruitment and data collection, were reviewed and approved by the university’s Institutional Review Board (IRB), ensuring that ethical standards for research involving human participants were upheld.

## 3. Results

This section presents findings from the post-survey and system usage data to evaluate student perceptions, experiences, and engagement with the Educational AI Hub. Key areas of analysis include how students compared the AI tool to traditional academic support, its perceived effectiveness across learning tasks, concerns about academic integrity, and attitudes toward ethical use and future integration of AI in education. The results reflect both the opportunities and challenges associated with implementing AI-powered tools in nursing education, providing insights into student trust, usability preferences, and expectations for responsible use.

### 3.1. Student Perceptions of AI Compared to Human Support

The post-usage survey included questions aimed at understanding how students evaluated the Educational AI Hub relative to more traditional sources of academic support, such as instructors, teaching assistants, or classmates. Students were asked to rate their experiences across three key areas: the helpfulness of the support provided, ease of access, and their level of comfort when seeking assistance. Each item was rated on a 5-point scale, ranging from “Much Worse” to “Much Better,” allowing for a comparative assessment of AI versus human support.

As shown in Figure 2, student perceptions of the Educational AI Hub compared to traditional academic support sources (such as instructors, teaching assistants, or peers) varied across three key dimensions: quality of help, convenience of access, and comfort in seeking assistance. In terms of convenience, the response was unequivocal, 100% of students rated the AI Hub as more convenient than human support. This suggests that the tool’s on-demand availability and seamless integration within the learning environment made it an especially attractive option for students seeking flexible, immediate assistance.

**Figure 2:**
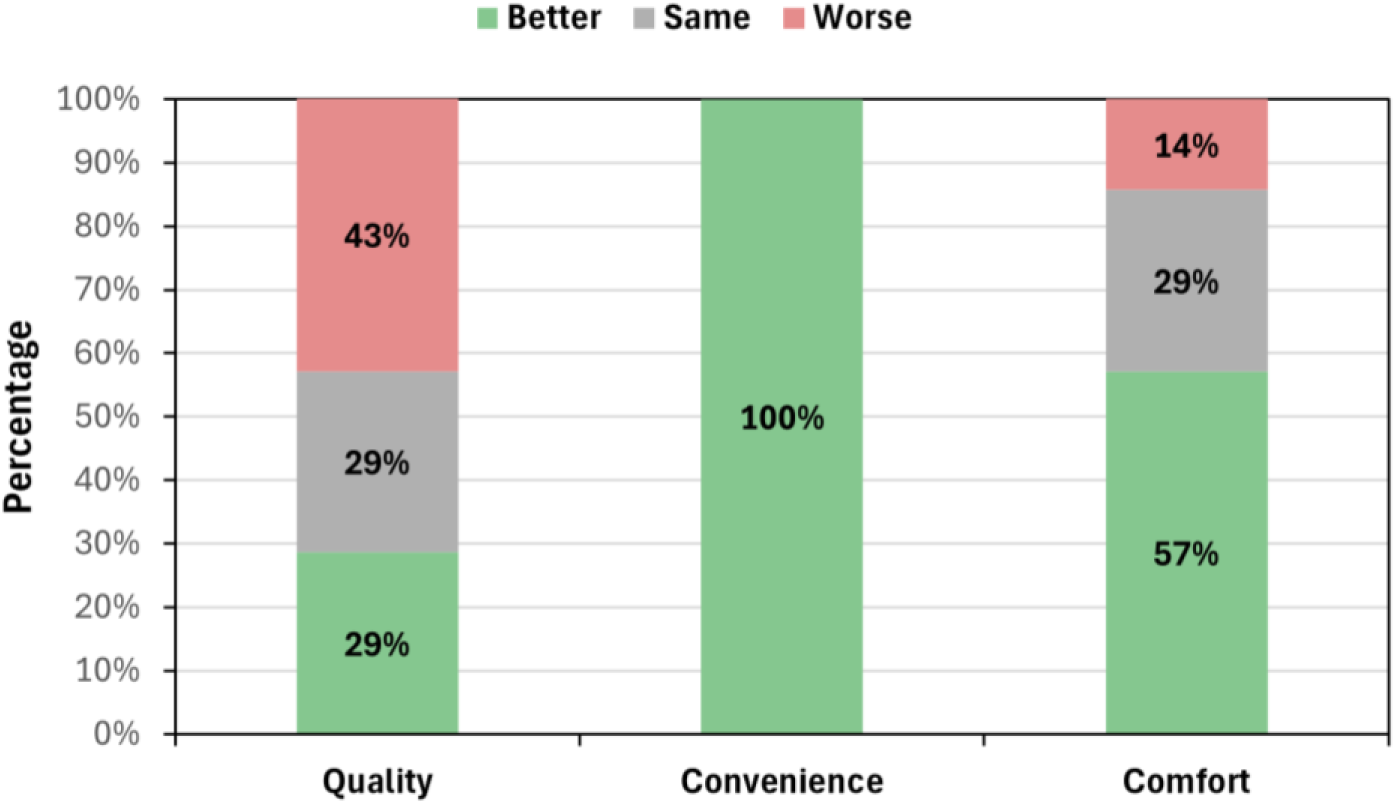
Student ratings of the AI Hub versus human support in terms of quality, convenience, and comfort (N=7)

Regarding comfort, a majority of students (57%) indicated they felt more comfortable using the AI tool than approaching a human source. An additional 29% reported no difference, while only 14% felt less comfortable. These results highlight the AI Hub’s potential to reduce social or performance-related anxieties that some students may experience when asking questions in traditional academic settings.

However, perceptions of help quality were more divided. While 29% of respondents found the AI Hub to offer better support than human counterparts, an equal percentage viewed it as comparable, and 43% rated it as worse. This indicates that while the tool may enhance access and comfort, it may not yet fully meet student expectations in terms of instructional depth or personalized academic guidance. These mixed ratings may reflect limitations in the AI’s ability to handle nuanced or highly contextual questions, especially in a complex discipline like nursing.

### 3.2. Barriers to Adoption and Ethical Concerns

Although students found the Educational AI Hub convenient and generally approachable, concerns related to trust and academic integrity emerged in the post-survey results. As shown in Figure 3, responses reflected varying degrees of skepticism and ethical apprehension regarding the use of AI in an academic setting.

**Figure 3:**
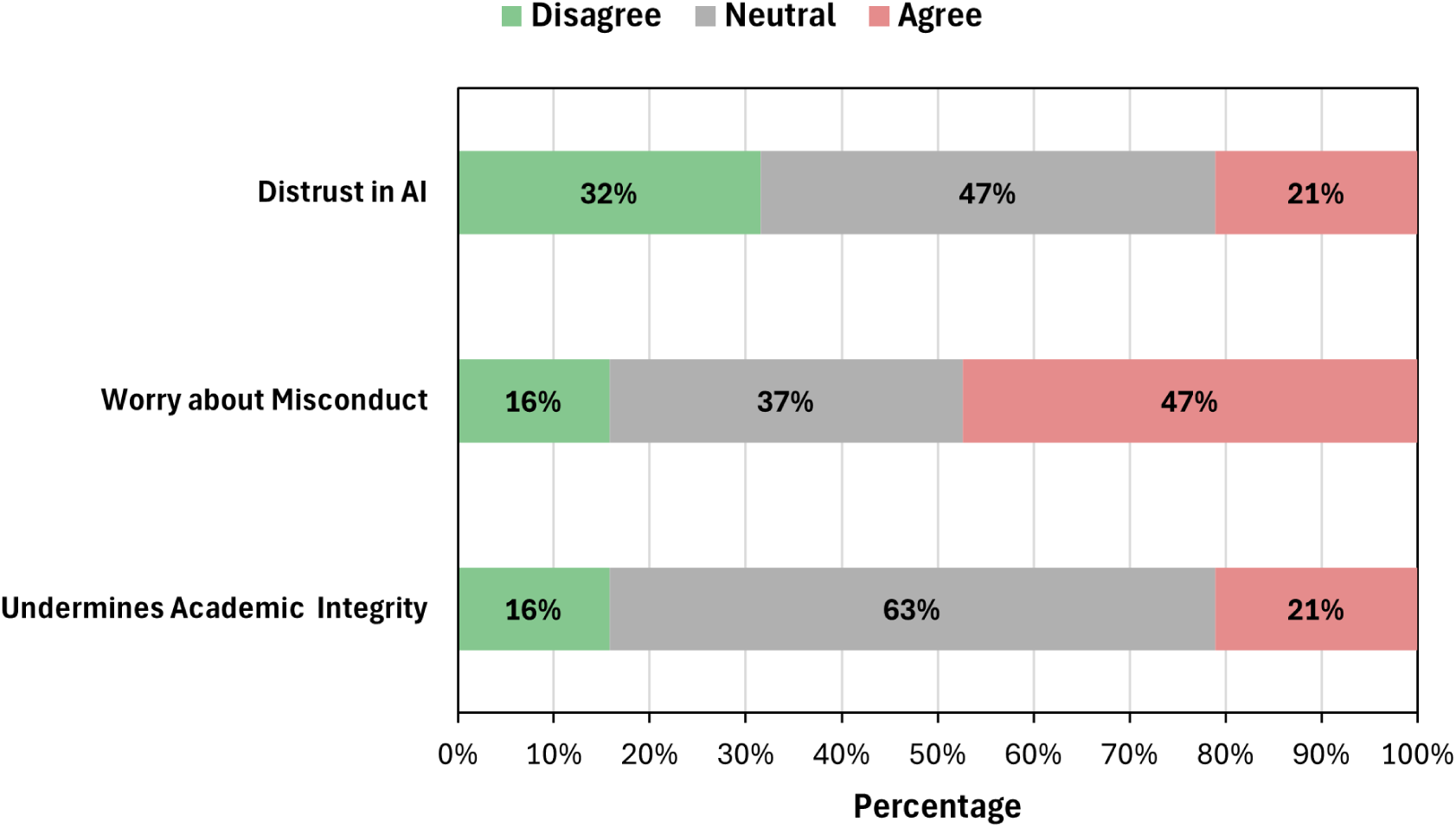
Student agreement levels on issues related to trust in AI, concerns about academic misconduct, and the potential impact of AI on academic integrity (N = 19)

Specifically, 21% of students expressed active distrust in the AI system, while nearly half (47%) remained neutral, suggesting a degree of uncertainty or caution about the tool’s reliability. Only 32% disagreed with the statement, indicating confidence in the tool. When it came to concerns about academic misconduct, 47% of students agreed that AI use could contribute to dishonest practices, while 37% were neutral, and just 16% disagreed. This highlights a clear ethical tension, with many students acknowledging the potential for misuse, even if they themselves were not engaging in it.

Finally, regarding the broader concern that AI may undermine academic integrity, only 21% agreed, while a large majority (63%) were neutral. This suggests that while students recognize risks, many are still forming their opinions or maybe awaiting institutional guidance on what constitutes appropriate AI use.

### 3.3. Student-Reported Usefulness Across Academic Tasks

Students rated the Educational AI Hub’s effectiveness across several areas of academic support, including understanding course material, studying, completing homework, improving grades, and contributing to overall learning. As presented in Figure 4, students expressed generally positive views, with the strongest endorsements seen for support with studying, concept comprehension, and overall learning.

**Figure 4:**
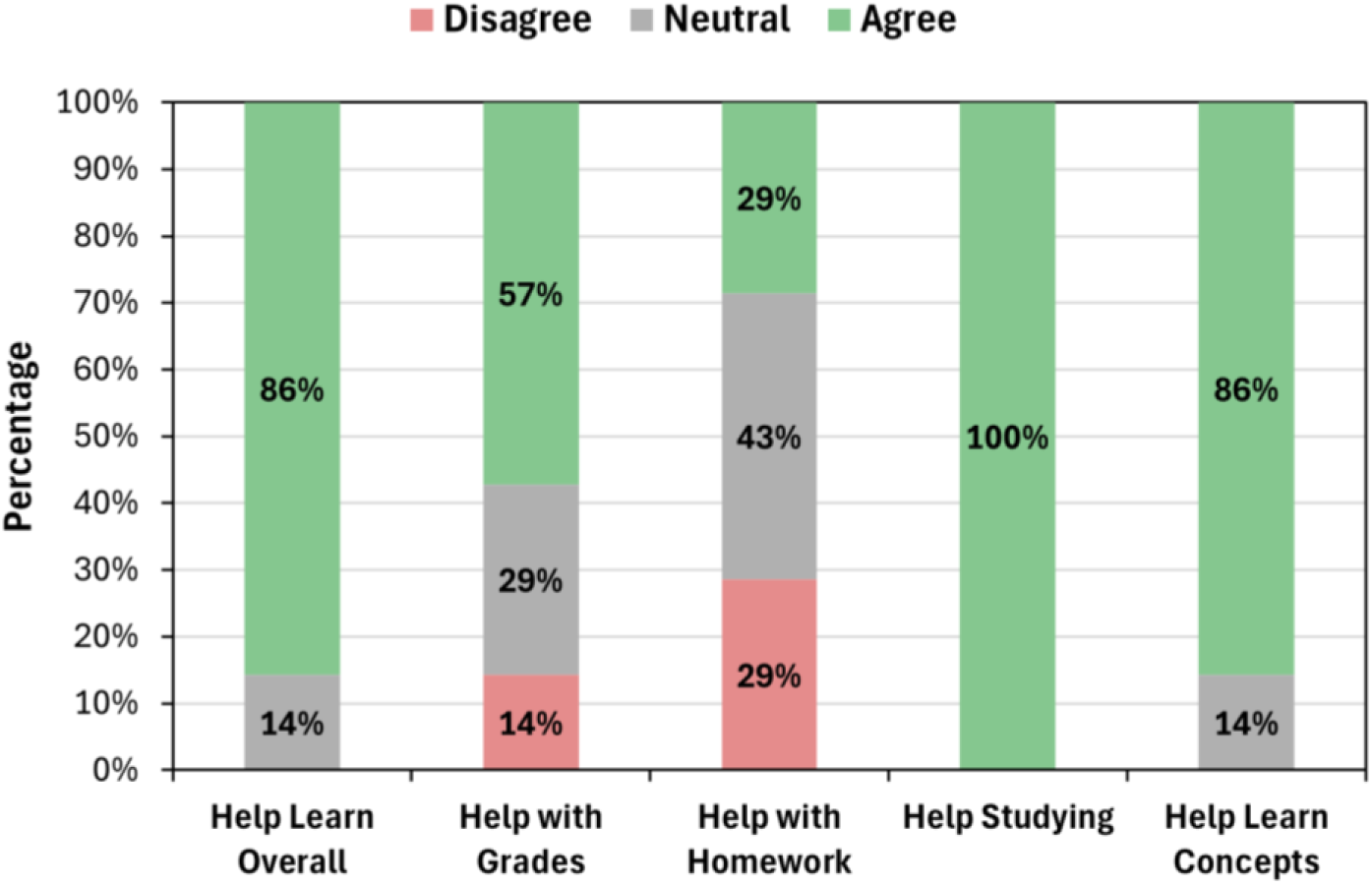
Reported usefulness of the Educational AI Hub across various academic tasks, as perceived by students (N = 7)

Specifically, 100% of students agreed that the AI Hub was helpful for studying, and 86% agreed it helped them both learn course concepts and improve overall learning. When asked about its impact on grades, 57% agreed it was beneficial, although 14% disagreed and 29% were neutral, suggesting that while many saw academic value, some remained unsure of its direct effect on performance metrics.

Perceptions of the tool’s usefulness for homework were more mixed. While 29% found it helpful, 43% were neutral and 29% disagreed, indicating that students may have used the AI Hub more for conceptual review than for task-specific assistance. These findings suggest that students most appreciated the AI Hub as a tool for understanding and reviewing material, rather than as a direct solution for assignments or grade improvement.

### 3.4. Student Certainty Around Misconduct and Views on AI Policy

In addition to exploring perceived usefulness and ethical concerns, the post-survey included questions about students’ certainty regarding what constitutes academic misconduct when using generative AI and their opinions on how strictly AI use should be regulated in course settings. As shown in Figure 5, most students expressed uncertainty about misconduct boundaries. While 58% of respondents said they were *somewhat* certain about what qualifies as academic misconduct with AI tools, only 11% felt *moderately* certain and just 5% reported being *very* certain. In contrast, 27% of students (16% *not certain*, 11% *a little certain*) lacked clarity entirely, indicating a need for clearer guidance and instruction around responsible AI use.

**Figure 5:**
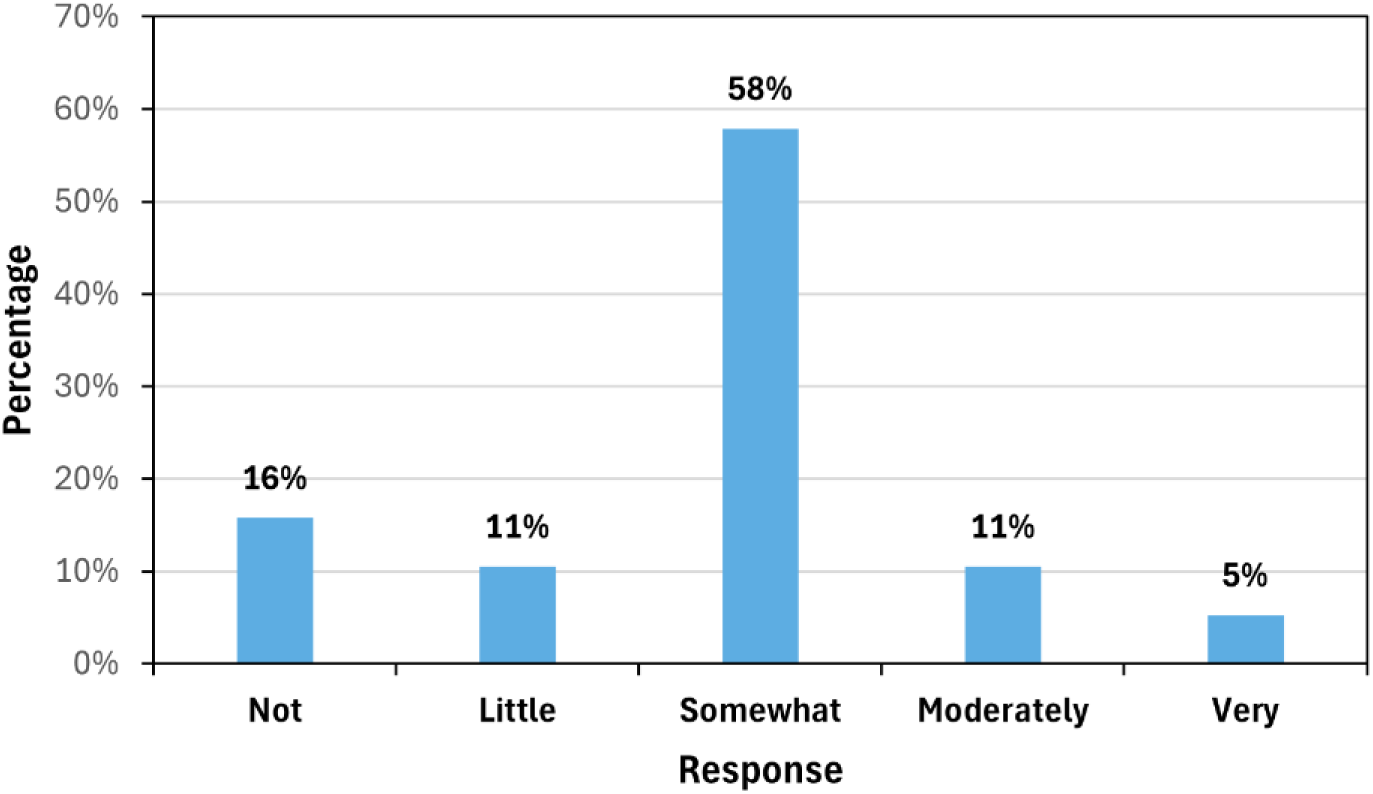
Student certainty about what constitutes academic misconduct when using AI tools (N = 19)

Figure 6 reflects students’ views on how much restriction should be placed on AI use in academic settings. A large majority, 84%, believed AI should be *somewhat* restricted, with 11% favoring *very* restrictive policies. Only a small number (5%) supported *less* restriction, and no respondents supported either *full bans* or *no restrictions* at all. These findings suggest that while students generally support access to AI tools, they also recognize the importance of boundaries to uphold academic integrity.

**Figure 6:**
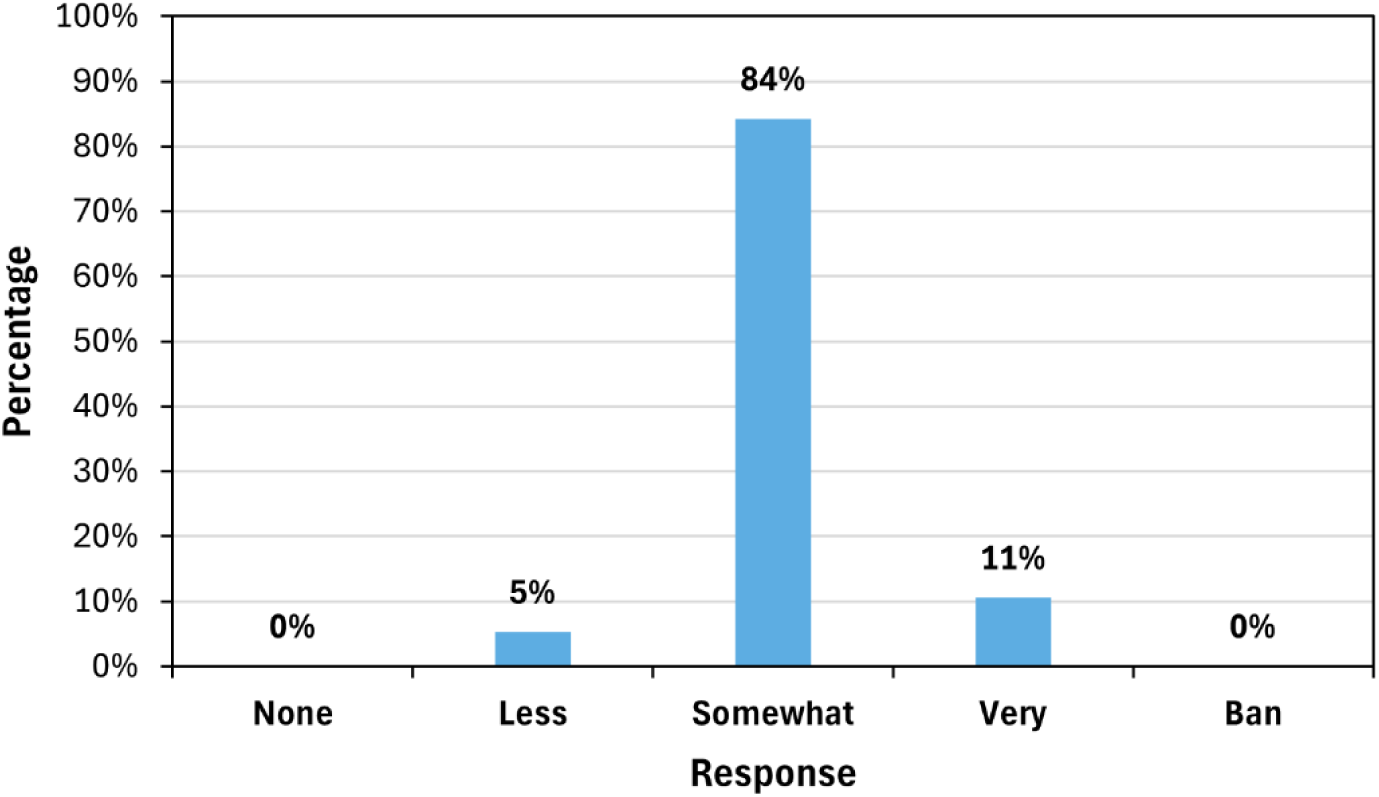
Student perspectives on how much AI use should be restricted in academic courses (N = 19)

### 3.5. Perceptions of AI as an Ethical Academic Aid and Future Use in Education

The post-survey also explored students’ views on the ethical acceptability of using AI tools in academic settings and their interest in seeing more AI integration in future courses. As illustrated in Figure 7, responses reflected a cautiously optimistic stance toward AI’s educational role. When asked whether using AI as an academic aid is ethical, 53% of students selected *neutral*, indicating uncertainty or a desire for clearer guidelines. However, 26% agreed with the ethical use of AI in coursework, while 21% disagreed, revealing a moderate split in opinion. In contrast, students expressed strong enthusiasm for expanding AI’s presence in education. Among those who responded to the item 100% indicated that they would like to see more AI tools integrated into their academic experiences, underscoring a growing interest in leveraging AI to enhance learning when implemented thoughtfully.

**Figure 7:**
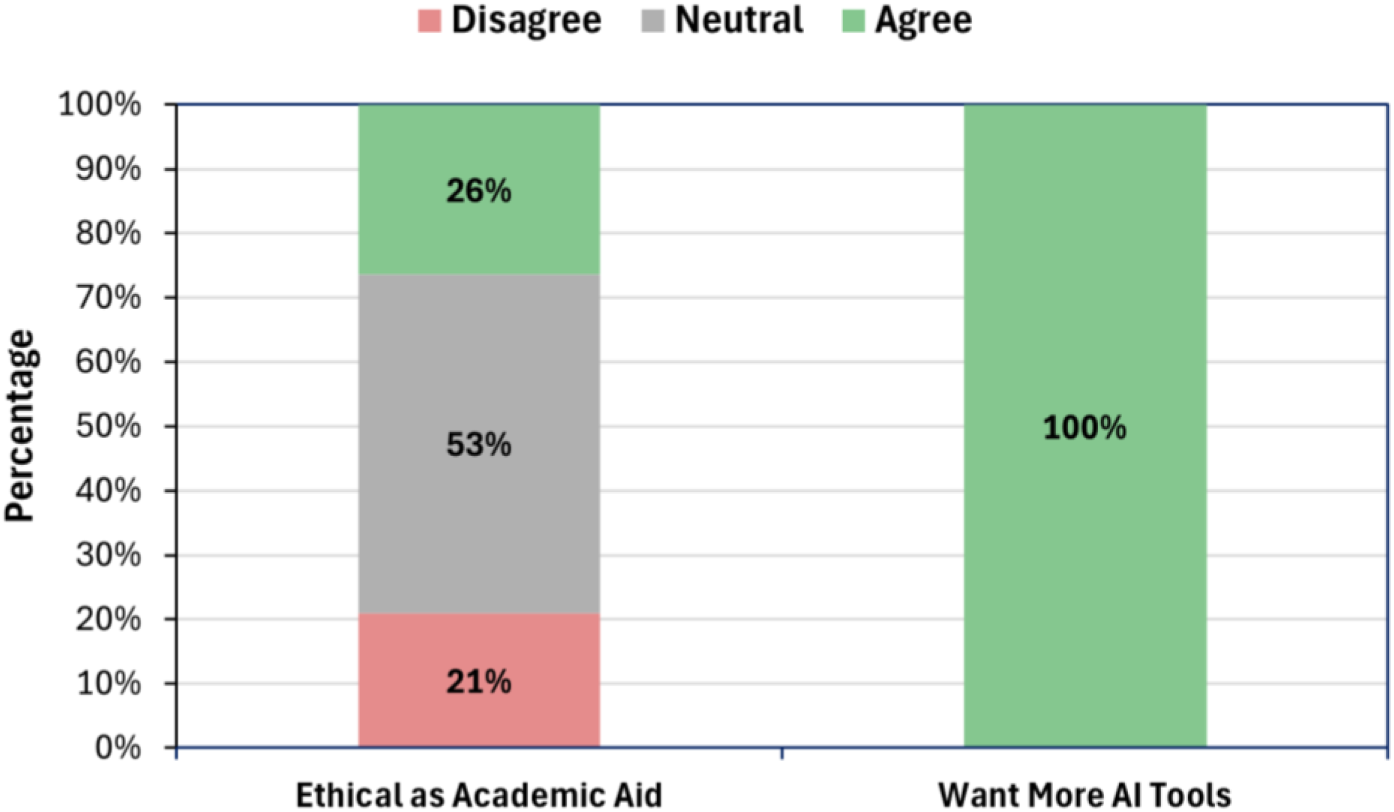
Student perceptions of the ethical use of AI in academics (N = 19) and interest in expanding the use of AI tools in future courses (N = 7)

## 4. Discussions

The results of this study indicate that nursing students found the Educational AI Hub to be a convenient and generally supportive academic tool, particularly when it came to accessing help quickly and independently. Students valued the tool’s 24/7 availability and its integration within the course’s learning environment, which made it especially useful for studying and reviewing course concepts. These findings align with previous research suggesting that AI-powered educational tools can enhance learner engagement, support personalized learning, and facilitate autonomous study (Srinivasan et al., 2023; Tam et al., 2023; Tseng et al., 2025).

However, while students reported high satisfaction with the tool’s accessibility and comfort, they expressed more varied opinions about the quality of academic support it provided. Nearly half of the respondents felt that the AI Hub was not as helpful as traditional human support, such as instructors or teaching assistants, especially for more complex or context-dependent questions. This indicates that while AI tools can effectively support surface-level learning tasks and immediate information needs, they may fall short in facilitating deeper, more nuanced forms of learning that require clinical reasoning, emotional nuance, or ethical judgment. These results support existing literature emphasizing the complementary, rather than substitutive, role of AI in health professions education.

A significant area of concern highlighted by this study was student uncertainty around the ethical use of AI tools. Many students reported a lack of clarity about what constitutes academic misconduct when using generative AI, and nearly half expressed concern that these tools could contribute to dishonest practices. While students generally did not perceive AI use as inherently unethical, the high level of neutrality and hesitation in their responses suggests a strong demand for clearer guidance. These findings highlight the need for institutions and educators to develop and communicate clear policies on acceptable AI use and to incorporate digital ethics education into nursing curricula. When students are unsure of the boundaries, they are more likely to use AI cautiously or avoid it altogether, even when it could enhance their learning.

Interestingly, while ethical uncertainty remains a concern, students showed a strong interest in the continued and expanded use of AI in their education. All students who responded to the question about future integration expressed a desire to see more AI tools incorporated into their learning environments. This enthusiasm underscores a general recognition of the value these tools can provide when thoughtfully implemented. Students appear to appreciate AI not as a replacement for human instruction but as a valuable supplement that enhances learning flexibility, particularly in resource-limited settings or outside traditional classroom hours.

The study offers several implications for nursing education. First, AI tools should be intentionally integrated into curricula as supplemental supports, especially in courses where independent study and content mastery are emphasized. Second, educators should provide clear guidance and explicit examples of what constitutes responsible versus inappropriate AI use, ideally embedded within discussions on academic integrity and professional ethics. Finally, institutions should take proactive steps to ensure that both students and faculty are equipped with the skills and knowledge needed to engage with AI critically and ethically.

Despite its contributions, this study is limited by a small sample size and its focus on a single course, which may limit generalizability. Additionally, the findings are based on self-reported perceptions and do not measure objective learning outcomes. Future research should explore how AI-enhanced tools affect academic performance, skill development, and long-term retention. It would also be valuable to investigate how perceptions differ across student levels, learning contexts, or types of AI tools.

## 5. Conclusion

This study examined nursing students’ engagement with the Educational AI Hub; an AI-powered academic support tool integrated into an undergraduate nursing course. The results demonstrate that students generally found the tool helpful, particularly for studying, reviewing course material, and accessing support outside of regular instructional hours. The AI Hub was perceived as highly convenient and comfortable to use, with many students appreciating its ability to provide immediate, personalized assistance in a self-paced format. These findings reinforce the growing potential of AI technologies to support independent learning and enhance educational access in nursing programs.

However, the study also uncovered significant ethical and pedagogical concerns. Many students expressed uncertainty about what constitutes appropriate use of AI in academic settings, particularly around issues of academic integrity and fairness. While few students viewed the use of AI as explicitly unethical, a majority were unsure, reflecting a broader lack of clarity around institutional expectations. Additionally, perceptions of the AI tool’s instructional quality were mixed, suggesting that students continue to rely on human educators for more complex, contextual, or emotionally nuanced guidance.

Despite these concerns, students overwhelmingly expressed a desire for more AI tools in future courses. This enthusiasm highlights a key opportunity for nursing education: to harness the strengths of AI while simultaneously addressing its limitations through clear policies, ethical frameworks, and thoughtful curricular integration. Faculty development, student training in digital literacy, and the inclusion of ethical discussions about AI use in coursework will be essential as the field moves forward.

As AI becomes increasingly embedded in educational and clinical environments, nurse educators are uniquely positioned to guide its responsible use. Tools like the Educational AI Hub, when implemented thoughtfully, can serve as valuable complements to traditional instruction, helping students build knowledge, confidence, and autonomy. To fully realize the benefits of AI in nursing education, institutions must invest in not just the technology itself, but the systems of support and accountability that surround it. With the right balance of innovation and ethical oversight, AI-enhanced tools can play a transformative role in preparing a competent, reflective, and future-ready nursing workforce.

## Data Availability

The datasets used and/or analyzed during the current study are available from the corresponding author on reasonable request.

## Acknowledgements

The authors would like to thank Jae-Eun Russell, Salim George, and Anna Smith for their invaluable support throughout the study. Their assistance with IRB approvals, coordination of study logistics, and thoughtful feedback during both the research and writing phases was instrumental to the success of this project. We gratefully acknowledge the Office of Teaching, Learning, and Technology (OTLT) at the University of Iowa for their guidance and support in implementing the Educational AI Hub and facilitating this evaluation.

## Funding

Funding for this project was provided by the University of Iowa’s Innovations in Teaching with Technology Awards.

## Ethics Approval and Consent to Participate

This study was approved by the Human Subjects Office Institutional Review Board (IRB), approval number 201707769. Informed consent was obtained from all participants prior to the administration of both pre- and post-surveys. Participation was entirely voluntary, and all individuals were fully informed about the purpose and procedures of the study. The authors affirm that this work was conducted in accordance with established ethical standards.

## Notes

### Competing Interest Statement

The authors have declared no competing interest.

### Author Declarations

The Institutional Review Board of the University of Iowa gave ethical approval for this work (approval number 201707769). Informed consent was obtained from all participants prior to the administration of both pre- and post-surveys. Participation was entirely voluntary, and all individuals were fully informed about the purpose and procedures of the study. The authors affirm that this work was conducted in accordance with established ethical standards.

## References

Abrar, M., Sermet, Y., & Demir, I. (2025). An empirical evaluation of large language models on consumer health questions. BioMedInformatics, 5(1), 12.

Ali, H., & Aysan, A. F. (2025). Ethical dimensions of generative AI: a cross-domain analysis using machine learning structural topic modeling. International Journal of Ethics and Systems, 41(1), 3–34.

Alshehri, F. D., Jones, S., & Harrison, D. (2023). The effectiveness of high-fidelity simulation on undergraduate nursing students’ clinical reasoning-related skills: A systematic review. Nurse Education Today, 121, 105679.

Baglivo, F., De Angelis, L., Casigliani, V., Arzilli, G., Privitera, G. P., & Rizzo, C. (2023). Exploring the possible use of AI chatbots in public health education: feasibility study. JMIR medical education, 9, e51421.

Balay-Odao, E. M., Omirzakova, D., Bolla, S. R., Almazan, J. U., & Cruz, J. P. (2024). Health professions students’ perceptions of artificial intelligence and its integration to health professions education and healthcare: a thematic analysis. Ai & Society, 1-11.

Benfatah, M., Youlyouz-Marfak, I., Saad, E., Hilali, A., Nejjari, C., & Marfak, A. (2024). Impact of artificial intelligence-enhanced debriefing on clinical skills development in nursing students: A comparative study. Teaching and Learning in Nursing, 19(3), e574–e579.

Bhutoria, A. (2022). Personalized education and Artificial Intelligence in the United States, China, and India: A systematic review using a Human-In-The-Loop model. Comput. Educ. Artif. Intell., 3, 100068. 10.1016/j.caeai.2022.100068

Castonguay, A., Farthing, P., Davies, S., Vogelsang, L., Kleib, M., Risling, T., & Green, N. (2023). Revolutionizing nursing education through AI integration: A reflection on the disruptive impact of ChatGPT. Nurse education today, 129, 105916.

Charow, R., Jeyakumar, T., Younus, S., Dolatabadi, E., Salhia, M., Al-Mouaswas, D., … & Wiljer, D. (2021). Artificial intelligence education programs for health care professionals: scoping review. JMIR Medical Education, 7(4), e31043.

Chi, N.C., Nguyen, K., Shanahan, A., Demir, I., Fu, Y.K., Chi, C.L., Perkhounkova, Y., Hein, M., Buckwalter, K., Wolf, M. and Williams, K., 2025. Usability Testing of the PACE-App to Support Family Caregivers in Managing Pain for People With Dementia. The Gerontologist, 65(2), p.gnae163.

De Gagne, J. C., Hwang, H., & Jung, D. (2024). Cyberethics in nursing education: Ethical implications of artificial intelligence. Nursing ethics, 31(6), 1021–1030.

Ghnemat, R., Shaout, A., & Al-Sowi, A. (2022). Higher Education Transformation for Artificial Intelligence Revolution: Transformation Framework. Int. J. Emerg. Technol. Learn., 17, 224–241. 10.3991/ijet.v17i19.33309

Han, S., Kang, H. S., Gimber, P., & Lim, S. (2025). Nursing Students’ Perceptions and Use of Generative Artificial Intelligence in Nursing Education. Nursing Reports, 15(2), 68.

Hawk, H., Coriasco, M., & Jones, J. R. (2024). Generative Artificial Intelligence: A Reflexive Thematic Analysis of Nursing Students’ Perceptions Following a Guided Learning Activity. Nurse educator, 10-1097.

Jallad, S. T., Alsaqer, K., Albadareen, B. I., & Al-Maghaireh, D. (2024). Artificial intelligence tools utilized in nursing education: Incidence and associated factors. Nurse education today, 142, 106355.

Kit, N., Luo, W., Chan, H., & Chu, S. (2022). An examination on primary students’ development in AI literacy through digital story writing. Computers and Education: Artificial Intelligence. 10.1016/j.caeai.2022.100054

Kizilkaya, D., Sajja, R., Sermet, Y., & Demir, I. (2025). Toward HydroLLM: a benchmark dataset for hydrology-specific knowledge assessment for large language models. Environmental Data Science, 4, e31.

Kowitlawakul, Y., Tan, J. J. M., Suebnukarn, S., Nguyen, H. D., Poo, D. C. C., Chai, J., … & Devi, K. (2022). Utilizing educational technology in enhancing undergraduate nursing students’ engagement and motivation: A scoping review. Journal of professional nursing, 42, 262–275.

Krive, J., Isola, M., Chang, L., Patel, T., Anderson, M., & Sreedhar, R. (2023). Grounded in reality: artificial intelligence in medical education. JAMIA Open, 6. 10.1093/jamiaopen/ooad037

Kwak, Y., Seo, Y. H., & Ahn, J. W. (2022). Nursing students’ intent to use AI-based healthcare technology: Path analysis using the unified theory of acceptance and use of technology. Nurse Education Today, 119, 105541.

Labrague, L. J., & Al Sabei, S. (2024). Integration of AI-Powered Chatbots in Nursing Education: A Scoping Review of Their Utilization, Outcomes, and Challenges. Teaching and Learning in Nursing.

Labrague, L. J., Aguilar-Rosales, R., Yboa, B. C., Sabio, J. B., & de Los Santos, J. A. (2023). Student nurses’ attitudes, perceived utilization, and intention to adopt artificial intelligence (AI) technology in nursing practice: a cross-sectional study. Nurse Education in Practice, 73, 103815.

Lane, S. H., Haley, T., & Brackney, D. E. (2024). Tool or tyrant: Guiding and guarding generative artificial intelligence use in nursing education. Creative Nursing, 30(2), 125–132.

Mady, A., & Niese, B. (2022). Augmenting AI and Human Capabilities in Competency-Based Learning. Proceedings of the 2022 *Computers and People Research Conference*. 10.1145/3510606.3550210

Montejo, L., Fenton, A., & Davis, G. (2024). Artificial intelligence (AI) applications in healthcare and considerations for nursing education. Nurse Education in Practice, 80, 104158.

Msambwa, M., Wen, Z., & Daniel, K. (2025). The Impact of AI on the Personal and Collaborative Learning Environments in Higher Education. European Journal of Education. 10.1111/ejed.12909

Ng, D., Leung, J., Su, J., Ng, R., & Chu, S. (2023). Teachers’ AI digital competencies and twenty-first century skills in the post-pandemic world. Educational Technology Research and Development, 71, 137–161. 10.1007/s11423-023-10203-6

Pursnani, V., Sermet, Y., & Demir, I. (2025). A conversational intelligent assistant for enhanced operational support in floodplain management with multimodal data. International Journal of Disaster Risk Reduction, 122, 105422.

Pursnani, V., Sermet, Y., Kurt, M., & Demir, I. (2023). Performance of ChatGPT on the US fundamentals of engineering exam: Comprehensive assessment of proficiency and potential implications for professional environmental engineering practice. Computers and Education: Artificial Intelligence, 5, 100183.

Saatçi, G., Korkut, S., & Ünsal, A. (2024). The effect of the use of artificial intelligence in the preparation of patient education materials by nursing students on the understandability, actionability and quality of the material: A randomized controlled trial. Nurse Education in Practice, 81, 104186.

Sajja, R., Pursnani, V., Sermet, Y., & Demir, I. (2025a). AI-assisted educational framework for floodplain manager certification: Enhancing vocational education and training through personalized learning. IEEE Access, 13, 42401–42413.

Sajja, R., Sermet, Y., & Demir, I. (2025b). End-to-end deployment of the educational AI hub for personalized learning and engagement: A case study on environmental science education. IEEE access.

Sajja, R., Sermet, Y., & Demir, I. (2025c). An Open-Source Dual-Loss Embedding Model for Semantic Retrieval in Higher Education. arXiv preprint arXiv:2505.04916.

Sajja, R., Sermet, Y., Cikmaz, M., Cwiertny, D., & Demir, I. (2024). Artificial intelligence-enabled intelligent assistant for personalized and adaptive learning in higher education. Information, 15(10), 596.

Sajja, R., Sermet, Y., Cwiertny, D., & Demir, I. (2023a). Platform-independent and curriculum-oriented intelligent assistant for higher education. International journal of educational technology in higher education, 20(1), 42.

Sajja, R., Sermet, Y., Cwiertny, D., & Demir, I. (2023b). Integrating AI and learning analytics for data-driven pedagogical decisions and personalized interventions in education. arXiv preprint arXiv:2312.09548.

Sajja, R., Sermet, Y., Fodale, B., & Demir, I. (2025d). Evaluating AI-powered learning assistants in engineering higher education: Student engagement, ethical challenges, and policy implications. arXiv preprint arXiv:2506.05699.

Sapci, A., & Sapci, H. (2020). Artificial Intelligence Education and Tools for Medical and Health Informatics Students: Systematic Review. JMIR Medical Education, 6. 10.2196/19285

Saritepeci, M., & Durak, H. (2024). Effectiveness of artificial intelligence integration in design-based learning on design thinking mindset, creative and reflective thinking skills: An experimental study. Educ. Inf. Technol., 29, 25175–25209. 10.1007/s10639-024-12829-2

Sermet, Y., & Demir, I. (2021). A semantic web framework for automated smart assistants: A case study for public health. Big Data and Cognitive Computing, 5(4), 57.

Shin, H., De Gagne, J. C., Kim, S. S., & Hong, M. (2023). The Impact of Artificial Intelligence-Assisted Learning on Nursing Students’ Ethical Decision-making and Clinical Reasoning in Pediatric Care: A Quasi-Experimental Study. CIN: Computers, Informatics, Nursing, 10-1097.

Silvestri-Elmore, A., & Burton, C. (2024). How Can Nursing Faculty Create Case Studies Using AI and Educational Technology?. Nurse Educator, 10-1097.

Simms, R. C. (2025). Generative artificial intelligence (AI) literacy in nursing education: A crucial call to action. Nurse Education Today, 146, 106544.

Srinivasan, M., Venugopal, A., Venkatesan, L., & Kumar, R. (2024). Navigating the pedagogical landscape: exploring the implications of AI and chatbots in nursing education. JMIR nursing, 7, e52105.

Sukmawati, I. D., Setyawati, M. B., & Prasetio, A. B. (2025). Gamification and ai in english language training for nurses: Trends and outcomes. In BIO Web of Conferences (Vol. 152, p. 01037). EDP Sciences.

Tam, W., Huynh, T., Tang, A., Luong, S., Khatri, Y., & Zhou, W. (2023). Nursing education in the age of artificial intelligence powered Chatbots (AI-Chatbots): are we ready yet?. Nurse Education Today, 129, 105917.

Topaz, M., Peltonen, L. M., Michalowski, M., Stiglic, G., Ronquillo, C., Pruinelli, L., … & Fukahori, H. (2024). The ChatGPT effect: nursing education and generative artificial intelligence. Journal of Nursing Education, 1-4.

Tseng, L. P., Huang, L. P., & Chen, W. R. (2025). Exploring artificial intelligence literacy and the use of ChatGPT and copilot in instruction on nursing academic report writing. Nurse Education Today, 147, 106570.

Xu, W., & Fan, O. (2021). A systematic review of AI role in the educational system based on a proposed conceptual framework. Education and Information Technologies, 27, 4195–4223. 10.1007/s10639-021-10774-y

Younas, M., El-Dakhs, D. A. S., & Jiang, Y. (2025). A comprehensive systematic review of ai-driven approaches to self-directed learning. IEEE Access.

Zhang, M., Zhu, L., Lin, S.Y., Herr, K., Chi, C.L., Demir, I., Dunn Lopez, K. and Chi, N.C., (2023). Using artificial intelligence to improve pain assessment and pain management: a scoping review. Journal of the American Medical Informatics Association, 30(3), pp.570–587.

Zhang, W., Cai, M., Lee, H. J., Evans, R., Zhu, C., & Ming, C. (2024). AI in Medical Education: Global situation, effects and challenges. Education and Information Technologies, 29(4), 4611–4633.

Zohny, H., McMillan, J., & King, M. (2023). Ethics of generative AI. Journal of medical ethics, 49(2), 79–80.

